# Pooling of Nasopharyngeal Swab Specimens for SARS-CoV-2 detection by RT-PCR

**DOI:** 10.1101/2020.04.22.20075598

**Authors:** Ignacio Torres, Eliseo Albert, David Navarro

## Abstract

Systematic testing of large population groups by RT-PCR is mandatory to Covid-19 case identification and contact tracing in order to minimize the likelyhood of resurgence in contagion. Sample pooling for RT-PCR has been effectively used to detect community transmission of SARS CoV-2. Nevertheless, this procedure may decrease the sensitivity of RT-PCR assays due to specimen dilution. We evaluated the efficacy of this strategy for diagnosis of Covid-19 using a sensitive commercially-available RT-PCR targeting SARS CoV-2 E and RdRp genes in a single reaction. A total of 20 mini-pools containing either 5 (n=10) or 10 (n=10) nasopharyngeal exudates collected in universal transport medium were made, each of which including a unique positive NP specimen. Positive specimens yielding C_T_ <32 for the E gene (6 out of 10) or <35.2 for the RdRP gene (7 out of 10) were detected in mini-pools of both sizes. In contrast, most NP samples displaying C_Ts_ > 35.8 for the E gene or 35.7 for the RdRP gene remained undetected in mini-pools of 5 specimens (3/4 and 2/3, respectively) or in mini-pools of 10 samples (4/4 and 3/3, respectively.

---

Until an effective vaccine is available, interruption of community circulation of SARS CoV-2 is crucial to control virus spread. To this end, systematic testing of large population groups by RT-PCR is mandatory to case identification and contact tracing thereby minimizing the likelyhood of resurgence in contagion.^1^ This approach faces a variety of obstacles, most notably the limited availability of reagents resources. Sample pooling for RT-PCR has been effectively used for screening of blood donors for Human immunodeficiency virus-1 and Hepatitis C virus in low-prevalence setttings.^2^ This strategy has also been applied to detect community transmission of SARS CoV-2 in the United States early in the pandemic, when virus circulation was low.^3^ Nevertheless, sample pooling may decrease the sensitivity of RT-PCR assays due to specimen dilution. Here, the efficacy of this strategy for diagnosis of Covid-19 was evaluated using a sensitive commercially-available RT-PCR (REALQUALITY RQ-2019-nCoV from AB ANALITICA; Padua, Italy, performed on the Applied Biosystems 7500 instrument), and low-size mini-pools. This RT-PCR assay targets the E (envelope) and RdRp (RNA dependent RNA polymerase) genes of SARS Cov-2 in a single reaction, with LODs of 125 and 150 copies/ml, respectively (according to the manufacturer). The current study was approved by the Ethics Committee of Hospital Clínico Universitario INCLIVA.

Nasopharyngeal specimens (NP) collected with flocked swabs in 3 ml of universal transport medium (Beckton Dickinson, Sparks, MD,USA), which had been stored at - 80°C, were retrieved for analysis. A total of 30 leftover specimens testing negative for SARS CoV-2 were mixed and used for pooling. In turn, 10 RT-PCR positive NP specimens yielding cycle threshold values (C_T_) ranging from 23.4 to 38.8 for the E gene, and 21.8 to 35.8 for the RdRP gene were selected for the experiments. A total of 20 mini-pools containing either 5 (n=10) or 10 (n=10) samples were made, each of which including a unique positive NP specimen (Table 1). These latter specimens had been collected from adult patients (median age, 61.5 years; range, 30-85) presenting with mild disease and not requiring hospitalization, at a median of 5 days (range, 1-7 days) following appearance of symptoms. Pertinent to this study is the fact that SARS CoV-2 load in upper respiratory tract specimens from symptomatic patients peaks within the first week after infection.^4,5^

**TABLE 1.** Detection of SARS CoV-19 RNA by RT-PCR in pooled nasopharyngeal specimens from patients with COVID-19.

| E (Envelope) gene |  |  | RpRd (RNA polymerase, RNA-dependent) gene |  |  |
| --- | --- | --- | --- | --- | --- |
| Non-pooled (C<sub>T</sub>) | Positive NP specimen pooled with 4 negative NP specimens (C<sub>T</sub>) | Positive NP specimen pooled with 9 negative NP specimens (C<sub>T</sub>) | Non-pooled (C<sub>T</sub>) | Positive specimen pooled with 4 negative NP specimens (C<sub>T</sub>) | Positive NPspecimen pooled with 9 negative NP specimens (C<sub>T</sub>) |
| 23.4 | 25.5 | 26.2 | 21.8 | 24.1 | 27.1 |
| 23.9 | 26.0 | 27.3 | 23.6 | 27.6 | 26.2 |
| 27.2 | 27.8 | 28.2 | 26.5 | 27.2 | 28.3 |
| 27.3 | 27.5 | 28.6 | 26.3 | 28.6 | 29.7 |
| 31.6 | 35.4 | 35.3 | 30.7 | 36.8 | 35.2 |
| 31.9 | 35.5 | 40.0 | 34.5 | 38.0 | 40.0 |
| 35.9 | ND | ND | 35.1 | 37.4 | 44.7 |
| 35.9 | ND | ND | 35.8 | ND | ND |
| 38.2 | ND | ND | 36.4 | ND | ND |
| 38.8 | 44.0 | ND | 35.2 | 43.4 | ND |

A volumen of 45 and 22.5 µL/specimen was used for preparing (manually) mini-pools of 5 and 10 samples, respectively, to achieve a final volumen of 225 µL, which was then mixed (1:1) with lysis buffer. RNA extraction was performed using the DSP virus Pathogen Minikit on the QiaSymphony Robot instrument (Qiagen, Valencia, CA, USA). RT-PCR was performed and interpreted according to the manufacturer instructions.
C<sub>T</sub>, cycle threshold; NP, nasopharyngeal exudate.

As shown in Table 1, positive NP specimens were detected in mini-pools of both sizes, as long undiluted samples yielded RT-PCR C_Ts_ <32 for the E gene (6 out of 10) or <35.2 for the RdRp gene (7 out of 10). As expected C_Ts_ were reached later in pooled samples. In contrast, most NP samples displaying RT-PCR C_Ts_ > 35.8 for the E gene or 35.7 for the RdRP gene remained undetected in mini-pools of 5 specimens (3/4 and 2/3, respectively) or in mini-pools of 10 samples (4/4 and 3/3, respectively).

Comprehensive testing policies in the community are primarily aimed at identifying SARS CoV-2-infected asymptomathic or paucisymptomatic individuals, who are known to represent a major source of transmission.^1^ Pooling strategies of RT-PCR testing may be advantageous when compared to assaying individuals samples separately if the proportion of positive specimens in the set of samples is low enough (∼1%)^6^. Previously reported data suggested that a single positive specimen could be detected in pools of up to 32 samples^5^, with a false negative rate of around 10%. In this work, pure RNA instead of original specimens were pooled for RT-PCR reactions.^6^ Likewise, identification of cases was achieved in pools of 48 samples including 1-5 RT-PCR positive samples. Our experience was less satisfactory, yet, positive specimens that went undetected when tested in pools presumably had very low viral loads, as inferred by RT-PCR CT values, most likely <6 log_10_ RNA copies/swab, which was shown to represent the viral RNA load threshold for virus infectivity^9^; thus, missing such type of positive results may have little consequences, if any, in terms of public health. It should mentioned that the efficiency of our approach may have been improved had fresh specimens been used for analyses, since degradation of RNA in samples during storage or thawing could have occurred. In summary, our data indicated that pooling NP specimens for RT-PCR testing may result in false negative results in patients presenting with mild Covid-19. The procedure evaluated herein can be improved by collecting samples in a smaller volumen of transport medium or increasing the number of RT-PCR cycles. Given the important benefit in terms of reagent savings inherent to this strategy, further studies are warranted to appraise its applicability in routine community surveys.

## Data Availability

Data available on request from the authors

## ACKNOWLEDGMENTS

No public or private funds were used for the current study. We are grateful to all personnel who work at Microbiology Service of Clinic University Hospital for their unconditional compromise in the fight against Covid-19.

## AUTHOR CONTRIBUTIONS

IT and EA performed the experiments and collected the data. DN analyzed the data and wrote the manuscript. All authors reviewed and approved the final version of the manuscript.

